# Polygenic Risk Scores Provide Strong Evidence for the Biologic Relevance of Molecular Subtypes of Schizophrenia

**DOI:** 10.1101/2024.06.21.24309320

**Authors:** C. Harker Rhodes, Richard Granger

**Author notes:** Contact Information: C. Harker Rhodes, MD, PhD 144, Sunset Rock Rd Lebanon, NH 03766.

## Abstract

We have previously described how the DLPFC transcriptomes of schizophrenia patients can be used to define two patient groups: “Type 1” patients with a relatively normal DLPFC transcriptome and “Type 2” patients with hundreds of genes differentially expressed in the DLPFC at a statistical significance which survives Bonferroni correction. The biological significance of the distinction between Type 1 and Type 2 patients remains an open question. In the present report we examine the polygenic risk scores (PRS) in those two groups of schizophrenic patients and observe Type1 and Type 2 schizophrenia have dramatically different genetic architectures. **That result supports the hypothesis that the DLPFC transcriptome-defined schizophrenia subtypes are biologically meaningful**.

There were three unexpected findings in this study.

The first unexpected finding was that PRS based on summary statistics from GWAS for schizophrenia captures very little of genetic contribution to the risk of Type 2 schizophrenia in the African-Americans in this study. Our working hypothesis is that in this Medical Examiner-derived cohort African-American ancestry may be a surrogate marker for socio-economic disadvantage and/or poor perinatal or obstetric care and that when present this environmental factor may overwhelm much of the genetic component for risk of type 2 schizophrenia. If in this particular cohort African-American ancestry is in fact a surrogate marker for socio-economic disadvantage, that makes this an especially valuable cohort for further study.

Of course, the fact that PRS based on any particular GWAS study does not capture the genetic risk for Type 2 schizophrenia in the African-Americans in this cohort does not imply that there is no genetic component to that risk. The second unexpected finding was that PRS for Cannabis Use Disorder captures genetic risk for Type 2, but not for Type 1 schizophrenia in the African-Americans in this cohort. The scientific and epidemiologic implications of this finding are unclear, but the finding rules out many of the technical artifacts that might have accounted for the failure of PRS based on the available GWAS studies of schizophrenia to capture that risk.

The third unexpected finding was that PRS based on summary statistics from GWAS for treatment-resistant schizophrenia captures genetic risk for Type 2, but not the Type 1 schizophrenia in the Caucasians in this cohort. **This is, perhaps, the most important finding in this study because it suggests that the distinction between Type 1 and Type 2 schizophenia is clinically as well as biologically meaningful**.

Disclosures: The opinions expressed herein are those of the authors and not necessarily representative of those of the Uniformed Services University of the Health Sciences (USU), the Department of Defense (DOD), the United States Army, Navy, Air Force, VA, NIH or any other US federal agency.

## Introduction

The popular “neurodevelopmental” hypothesis for the pathogenesis of schizophrenia postulates that schizophrenia is the result of some early pathogenic event occurring around the time of birth which predisposes the patient to the disease without producing clinical symptoms, and that the early event is followed by later event(s) during adolescence which trigger the appearance of clinical symptoms (see, for example, Weinberger et al., 1996). While the early and late events are presumably both due to a combination of environmental and genetic factors, the specific environmental and genetic factors are almost certainly different for the two events.

An alternate formulation of the neurodevelopmental hypothesis views schizophrenia as a syndrome, not a single disease. According to this alternate hypothesis there are two distinct diseases which can produce the schizophrenic phenotype, one of which is a result of perinatal event(s) while a second results from pathogenic event(s) in adolescence. Just as dementia often results from a combination of Alzheimer’s disease and vascular dementia, so the schizophrenic phenotype often results from the comorbid presence of both forms of schizophrenia. This formulation of the neurodevelopmental hypothesis predicts that schizophrenic patients lie on a biologic spectrum with some patients for whom the schizophrenic phenotype is due primarily to the perinatal event, while for other patients it is the primarily the adolescent event which is driving the disease pathogenesis. Presumably the gene x environment interactions are different for these two pathogenic events and many of the genetic risk alleles for schizophrenia play a role in the pathophysiology of either the early event or the late event, but not both.

The present study builds on observation that schizophrenic patients can be divided into two groups based on the extent to which their DLPFC transcriptomes are abnormal (Bowen et al. 2019). That observation was replicated at the protein level by autoradiographic measurement of the expression of the biologically important sphingosine phosphate receptor in the DLPFC of those patients (Chand et al. 2022). In other words, there appear to be two biologically distinct groups of schizophrenic patients; the extent to which this distinction represents a dichotomy or a continuum is unknown.

Genome-wide studies of large cohorts have identified many genetic polymorphisms associated with an increased risk of schizophrenia, and polygenic risk scores (PRS) based on those studies have been shown to account for around 15% of the variance in risk for schizophrenia (see, for example of the Psychiatric Genetics Consortium wave 3 results (Trubetskoy et al. 2022)). Presumably those PRS include both polymorphisms affecting the consequences of the perinatal event and polymorphisms affecting the neurologic functioning of the adult brain. The formulation of the neurodevelopmental hypothesis outlined above predicts that different polymorphisms are important for Type 1 vs Type 2 schizophrenia and PRS based on a variety publicly available GWAS summary statistics should distinguish the two diseases. This study tests that prediction.

## Materials and Methods

### The cohort on which this study is based

This study is a re-analysis of the data from multiple, largely overlapping cohorts consisting of schizophenic patients and neurotypical controls whose postmortem brains are presently available in the brain banks operated by the Human Brain Collection Core (HBCC) of the NIMH intramural research program and by the Lieber Institute for Brain Development (LIBD). These brains were initially collected by the staff of the Clinical Brain Disorders Branch (CBDB) of the NIMH intramural program and the collection further expanded by both the HBCC and the LIBD after Dr. Weinberger and many of the members of the CBDB moved to the Lieber Institute. At the time the Weinberger lab moved to the Lieber institute the cerebrum of many of the brains were split in the midsagital plane with one hemisphere remaining at the NIMH and the other half taken to the Lieber Institute.

We will refer to the subjects whose brains were collected before the Weinberger lab moved from NIMH to the Lieber Institute as the “CBCB cohort”. Subjects in the CBDB cohort for whom one hemisphere was taken to the Lieber institute as well as subjects whose brains were collected by the LIBD will be referred to as the “LIBD cohort”. Subjects in the CBDB cohort as well as those whose brains were collected by the HBCC after the departure of Dr. Weinberger will be referred to as the “HBCC cohort”. For the purposes of the present study the CBDB cohort was restricted to Caucasian or African-American adult schizophrenic patients and neurotypical controls for whom genome-wide genetic data is available. The subjects in the CBCB/HBCC/LIBD cohorts each have a unique identifier, the “Brain Number” which allows data collected by all of those laboratories to be combined in the present study.

The brains in the HBCC and LIBD brain banks were, with appropriate consent from the next of kin, provided to the CBCB/HBCC/LIBD by several Medical Examiners in the Washington, DC area.

Because medical examiners do not in general take jurisdiction of cases where the patient died in a hospital or under the care of a physician, this cohort of convenience is almost certainly not representative of the general population. As might be expected in a Medical Examiner cohort where the control subjects include accidental death and homocide victims, men are over-represented in the neurotypical controls. For some reason that imbalance is much more prominent in the Caucasians than African Americans. We speculate without evidence individuals of lower socio-economic status are almost certainly over represented in this cohort compared to the general population.

### Gene expression and genetic data

This study is an extension of the study reported by Bowen et al. (2019) in which expression array data from the dorsolateral prefrontal cortex (DLPFC) of subjects in the CBDB cohort was used to classify the schizophrenic patients as either “Type 1” or “Type 2”. That expression array data is publicly available from the NIMH Data Archive (NDA).

The array-based genotyping of these subjects, the quality control of that data, and the genome-wide imputation to GRCh37/hg19 is described by Duncan et al (2023). The data was provided to us by the HBCC under a data transfer agreement, but it is also publicly available from the NDA.

### GWAS summary statistics on which this study is based

#### Psychiatric Genetics Consortium schizophrenia wave 3 core dataset (Trubetskoy et al. 2022)

This monumental GWAS study was based on a 90 cohorts of European (EUR) and East Asian (ASN) ancestry containing a total of 67,390 schizophrenic patients and 94,015 neurotypical controls. This combined “core” cohort is composed primarily of Caucasians with about 13% Asians. It contains no Africans or African Americans. The GWAS summary statistics are contained in the file “PGC3_SCZ_wave3.core.autosome.public.v3.vcf.tsv.gz” which was downloaded from https://figshare.com/articles/dataset/scz2022/19426775

#### Psychiatric Genetics Consortium schizophrenia African-American cohort (Trubetskoy et al. 2022)

In addition to the “core” dataset, the Psychiatric Genetics Consortium wave 3 data includes summary statistics from 9 cohorts containing a total of 6152 African-American schizophrenic patients and 3918 African-American neurotypic controls. Those summary statistics are contained in the file “PGC3_SCZ_wave3.afram.autosome.public.v3.vcf.tsv.gz” which was also downloaded from https://figshare.com/articles/dataset/scz2022/19426775 For additional information about the GWAS study of those African-American schizophrenic patients and controls, see Bigdeli et al., 2020.

#### Cannabis Use Disorder GWAS based on multiple very large cohorts (Levey et al. 2023)

PRS for cannabis use disorder (CUD) is based on the summary statistics for the Caucasians in this very large, multicenter study which includes subjects in the Million Veteran Program (Hunter-Zinck et al. 2020). The summary statistics were downloaded from https://medicine.yale.edu/lab/gelernter/stats/#project-4 on 01/04/2025.

#### Treatment Resistant Schizophrenia

PRS for treatment resistant schizophrenia (TRS) was based on GWAS summary statistics estimated by Pardiñas et al., 2022 using the method suggested by Altman and Bland (2003) to calculate the genome-wide difference between two GWAS studies. Those authors used a clinical history of clozapine treatment as a surrogate for TRS the CLOZUK1 and CLOZUK2 cohorts for their GWAS data of individuals with TRS, giving them a combined cohort of 10,501 TRS patients and 24,542 neurotypical controls. For their non-TRS schizophrenic cohort they used the PRS wave 3 meta-cohort, removing patients with a history of clozapine treatment from those cohorts where that information was available and, for the cohorts where clozapine treatment history was not available, taking the conservative approach of assuming none of the patients had TRS. They then estimated a TRS-specific regression β coefficient for each SNP as the difference between the log(odds ratio) in the two studies and the standard error as the root mean squared of the standard errors of the two odds ratios. The resulting effect size was transformed to a z-score and used to estimate the P-value. The summary statistics from that imputed-GWAS were downloaded from https://walters.psycm.cf.ac.uk/ on 2/16/2025.

Important note: The PRS calculated based on this imputed-GWAS data is not PRS for treatment resistant schizophrenia. It is rather a polygenic score for the genetic risk factors which differentiate TRS from non-TRS.

### Calculation of PRS statistics

PRS statistics were calculated using the clumping and thresholding approach as implemented in the PRSice software package (Choi and O’Reilly 2019)^1^. The varience in risk attributable to PRS was estimated based on Nagelkerke’s R squared calculated using the R function Nagelkirke in the package {fsmb} (https://search.r-project.org/CRAN/refmans/fmsb/html/Nagelkerke.html).

## Results

### Cohort demographics

This cohort is a convenience sample based on Medical Examiner cases for whom the next of kin consented to the use of post-mortem tissue for this purpose. It is, therefore, not necessarily representative of the general population and this study limitation needs to be kept in mind when interpreting these results.

There were six self-reported Caucasians and four self-reported African Americans whose genetically imputed ancestry differed from their self-reported ancestry; those subjects were excluded from further analysis. Because there were so few individuals with self-reported Hispanic or Asian ancestry they were also excluded from further analysis. The final cohort is composed of individuals whose self-reported and genetically imputed ancestry is concordant and is either Caucasian or African American. The final cohort is well balanced for diagnosis and race (table 1). Additional cohort demographics are described in Bowen et al., 2019.

**Table 1:**
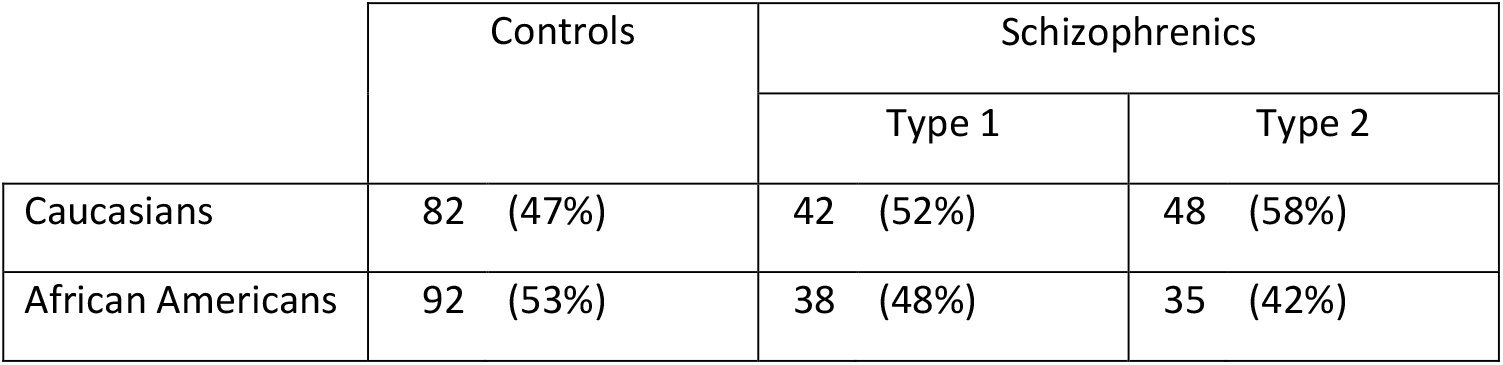
Distribution of cohort subjects by ancestry and diagnosis. Most importantly, the percentages of Type 1 and Type 2 patients is the roughly same in the Caucasians and in the African Americans.

|  | Controls | Schizophrenics |  |
| --- | --- | --- | --- |
|  |  | Type 1 | Type 2 |
| Caucasians | 82 (47%) | 42 (52%) | 48 (58%) |
| African Americans | 92 (53%) | 38 (48%) | 35 (42%) |

**Table 1:**
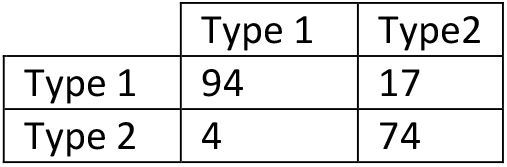
Contingency table showing the number of schizophrenic patients classified as Type 1 or Type 2 using the original WGCNA-based algorithm in Bowen et al., 2019 (columns) or the more appropriate algorithm developed by Kimes et al., 2017 (rows).

### Replication of clustering of schizophrenic patients using a more appropriate clustering algorithm

In our previous study (Bowen et al. 2019), WGCNA was used to cluster the schizophrenic patients based on the similarity of their DLPFC transcriptomes. An important limitation of that study is that the heirarchical clustering algorithm use in WGCNA is a greedy algorithm which will identify clusters (“modules”) without regard to whether or not that clustering is statistically significant. In other words, if subjects are clustered based on data which is a combination of some biologically relevant measurement and noise in that measurement, no statistical testing is done to see whether the clusters identified are the result of the biologic signal or the noise^2^. We therefore replicated the clustering of the schizophrenic patients using the Significance of Hierarchical Clustering (SHC) algorithm, a hierarchical clustering algorithm developed by Kimes et al., 2017 which tests the statistical significance of the clusters identified and only reports those with a family-wise error rate of less than 0.05.

The underlying assumption of the SHC algorithm is that the data is sampled from an independent multivariate Gaussian distribution. Since expression data of multiple genes is neither independent nor gaussian we first used principle component analysis to generate an independent multivariate distribution and then transformed the data to an independent multivariate Gaussian distribution using an inverse hyperbolic sine transformation^3^.

Using the transformed principle components of the expression array data not only allowed us to calculate the statistical significance of the hierarchical clustering of the patients (figure 1B), but it also provided a nice graphical illustration of the fact that the distribution of patients based on this metric is, in fact, bimodal (figure 1A).

**Figure 1A:**
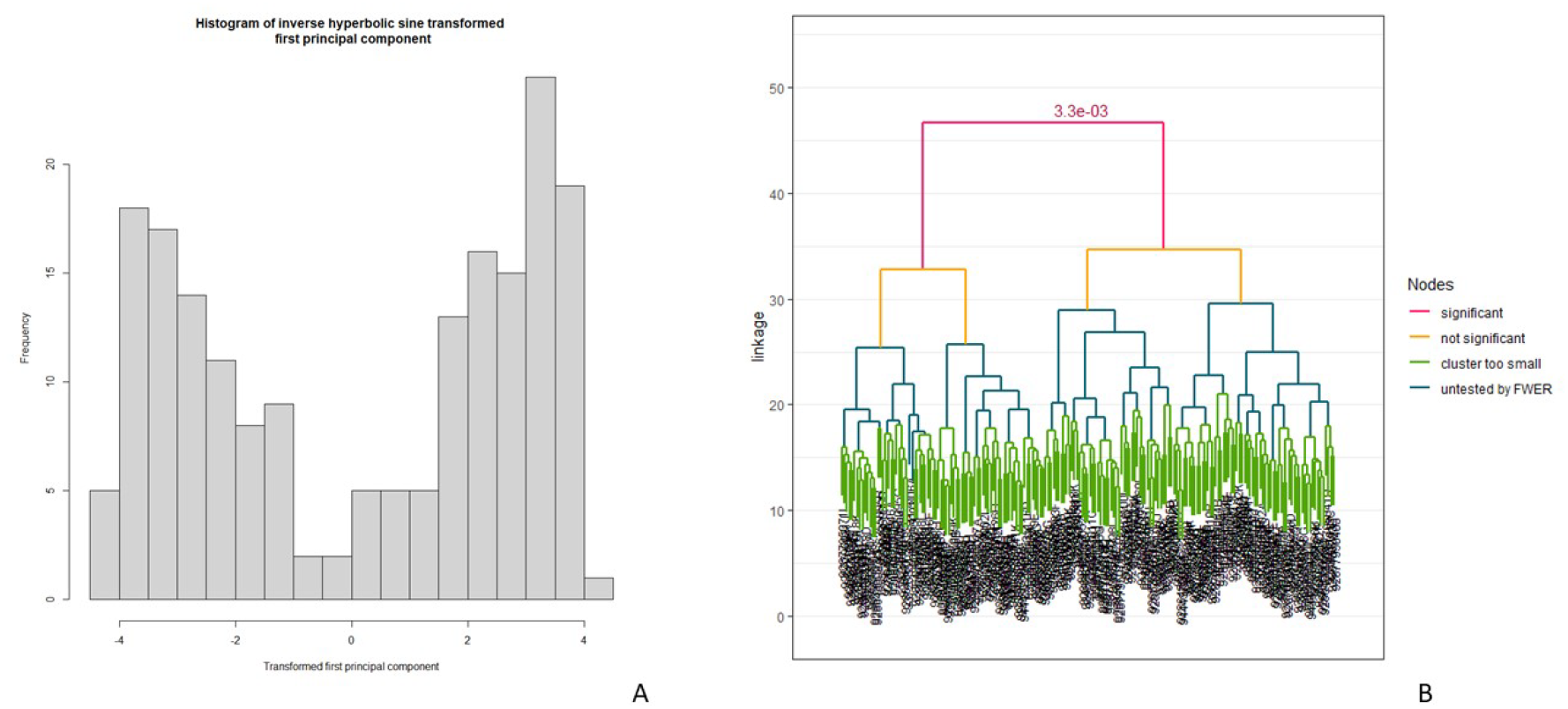
Histogram of the inverse hyperbolic sine transformed first principle component of the expression array data of the schizophrenic patients. Note the unequivally bimodal nature of the distribution, consistent with the hypothesis that these schizophrenic patients are drawn from a population containing patient two biologically distinct diseases. Figure 1B: Clustering of the schizophrenic patients using the Significance of Hierarchical Clustering (SHC) algorithm (Kimes et al. 2017). The clusters are deemed to be significant if the family-wise error rate (FWER) was less than 0.05

As can be seen in table 1, there was excellent, but not perfect agreement in the classification of the schizophrenic patients as either Type 1 or Type 2 done this way or as originally reported in (Bowen et al. 2019)

### There is a different genetic architecture of type 1 and type 2 schizophrenia

Figure 2 shows the variance in the risk of schizophrenia attributable to the polygenic risk score based on the summary statistics from the Psychiatric Genetics Consortium wave 3 core cohort GWAS study (Trubetskoy et al. 2022). In that figure the panels enclosed by the blue box show the results when the schizophrenic patients are pooled without regard to disease subtype. Those results are included only for comparison with other published studies which do not take disease subtype into account. The panels enclosed by the green box show the results when the subjects are pooled without regard to ancestry. Those results are included only for comparison with other published studies which do not take ancestry into account. The novel, scientifically important observations are illustrated in the four panels in the lower right of the figure enclosed by the red box.

**Figure 2:**
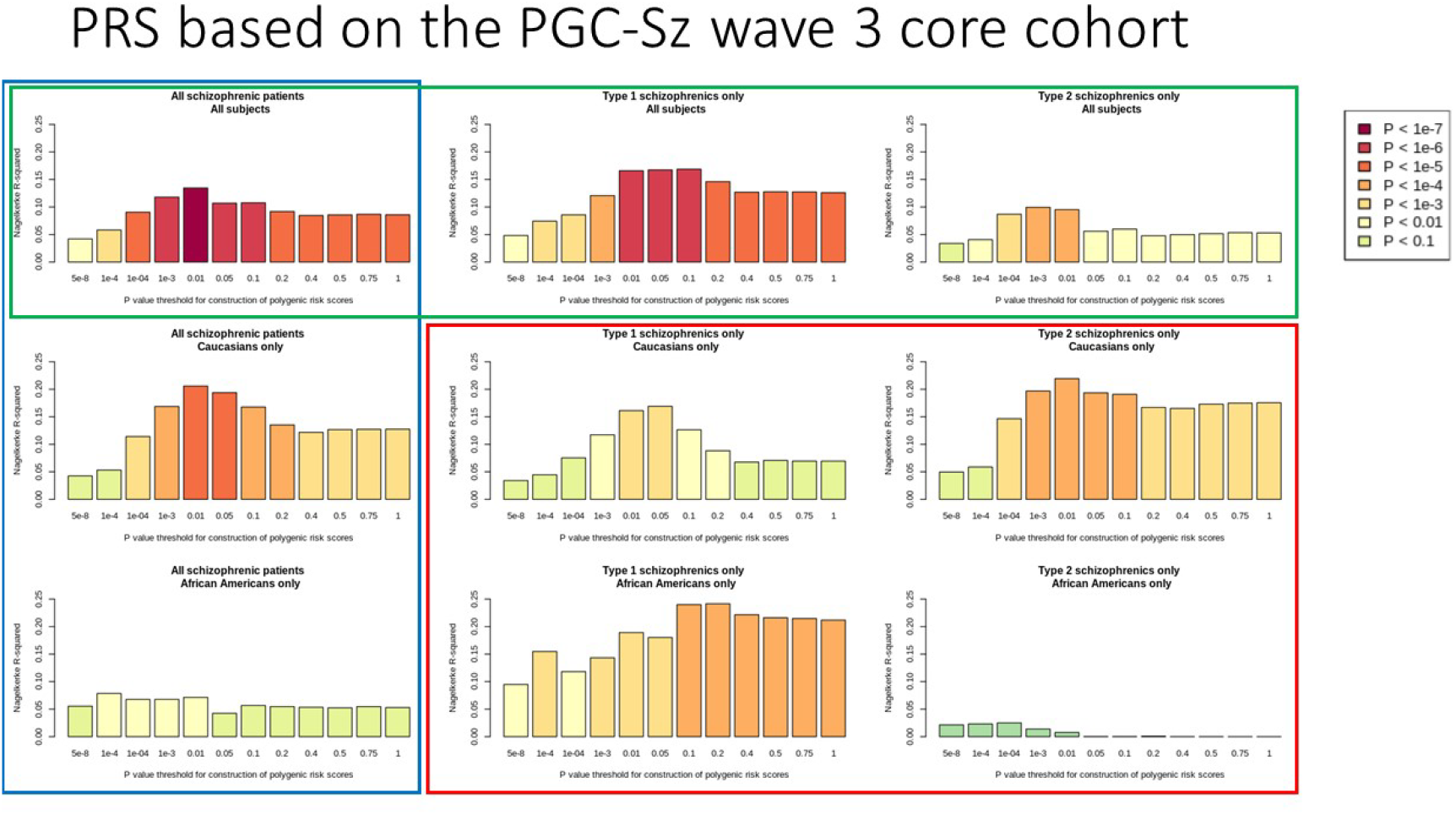
The fraction of variance in schizophrenia risk accounted for by PRS based on the Psychiatric Genetics Consortium wave 3 cohort (Trubetskoy et al. 2022).

The ordinate (the bar heights) is Nagelkirke’s pseudo-R^2^, a measure of the fraction of the variance accounted for by PRS. The statistial significancs of the logisitic regression on which the Nagelkirke’s pseudo-R^2^ is based is indicated by the bar colors.

Focusing on the four panels in the of red box figure 2, it is interesting to note that in the Caucasian subcohort, there appears to be a difference in the genetic architecture of Type 1 as compared to Type 2 schizophrenia when the polymorphisms with relatively low statistical significance (P-threshold > 0.2) are included in the PRS. A much more striking observation however, is that in the African-American subcohort virtually none of the variance in risk of Type 2 schizophrenia is accounted for by the PRS. In other words, based on this specific PRS, Type 1 and Type 2 schizophrenia clearly have a different genetic architecture in African-Americans. **The fact that any PRS reveals this distinction in any ancestry cohort is very strong evidence for a fundamental biologic difference between Type 1 and Type 2 schizophrenia**.

### Replication using PRS based on GWAS summary statistics from an African-American cohort

When evaluating the results illustrated in figure 1, it is important to remember that the cohort for the GWAS summary statistics on which that PRS is based (the PGC wave 3 core cohort) is composed primarily of Caucasians with about 13% Asians. It contains no Africans or African Americans. As a result, that PRS may fail to fully capture the genetic risk of schizophrenia in African-Americans because of differences between Caucasians and African-Americans in minor allele frequency, linkage disequilibrium, or some other genetic statistic.

We therefore repeated the analysis illustrated in figure 1 using PRS based on the summary statistics from the GWAS study of the Psychiatric Genetics Consortium wave 3 African American cohort (figure 3). The African-American cohort on which those PRS statistics are based is only about 10% the size of the Psychiatric Genetics Consortium “core” cohort and as a result the African-American GWAS based PRS statistics account for much less of the variance in risk of schizophrenia than the corresponding Caucasian GWAS based PRS statistics illustrated in figure 1. However, as illustrated by the panels in the red box, the important observation remains unchanged – apparently environmental factors play such an important role in the risk of Type 2 schizophrenia in African American patients that they overwhelm the genetic contribution to that risk which is captured in the PGC study. **This does not imply that there is no genetic component to this risk, only that it was not captured in the PGC study which did not take into account the existence of these subtypes**.

**Figure 3:**
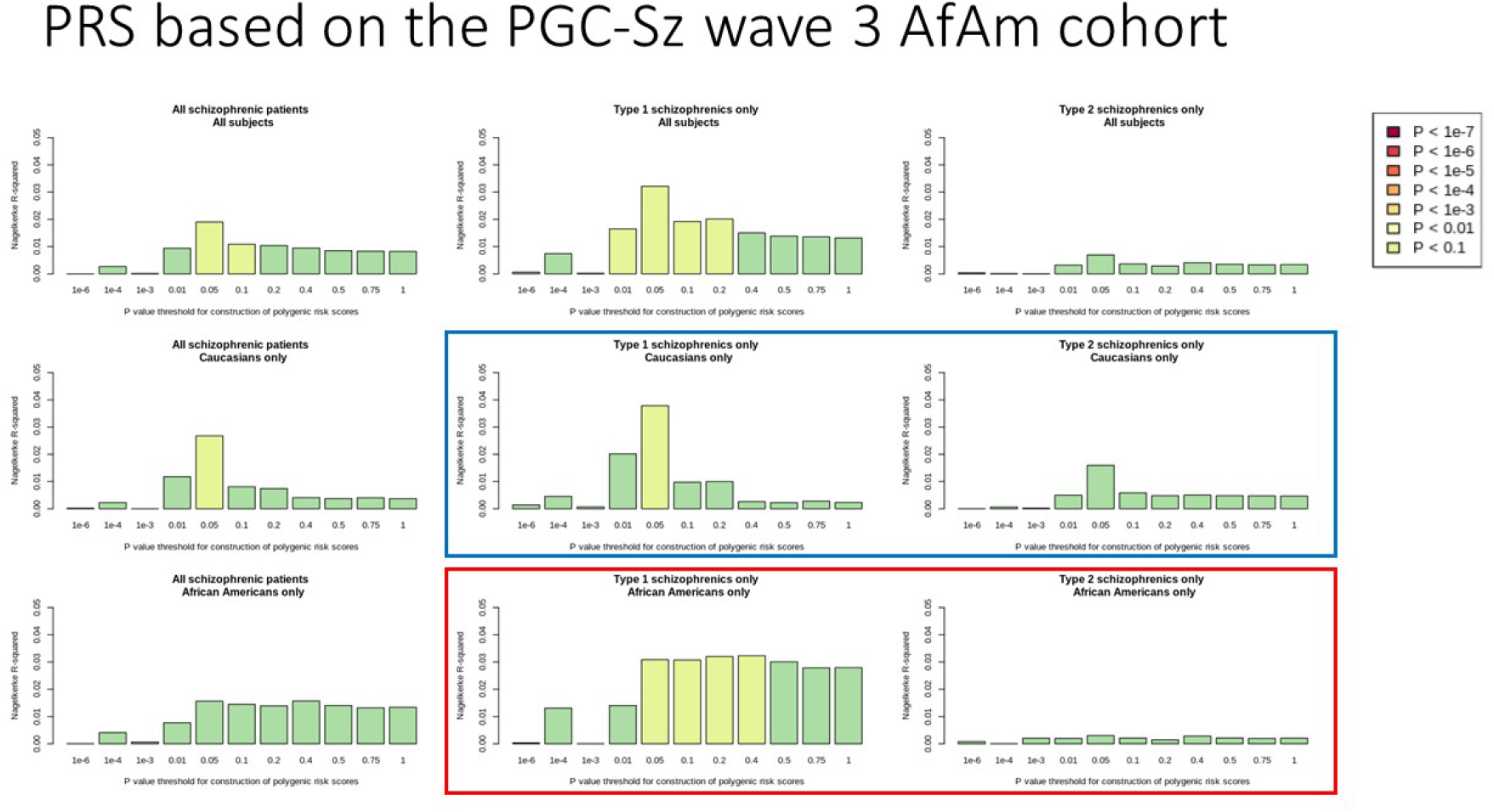
The fraction of variance in schizophrenia risk accounted for by PRS based on the Psychiatric Genetics Consortium wave 3 African-American cohort (Trubetskoy et al. 2022).

An important corollary of that conclusion is that if little or no genetic contribution to the risk of Type 2 schizophrenia in African American patients in GWAS studies which pool Type 1 and Type 2 schizophrenic patients, then this GWAS study of schizophrenia in African-Americans will be expected to capture only genetic features associated with the risk of Type 1 schizophrenia but not of Type 2 schizophrenia in Caucasian patients. That is in fact what is seen when the summary statistics from the African-American GWAS study are used to calculate PRS-associated risk of schizophrenia in Caucasian patients (the blue box in figure 3).

### A serendipitous identification of a PRS which captures some of the genetic risk for Type 2 schizophrenia in the African-Americans in this cohort

Cross-diagnostic PRS (the use of PRS based on GWAS for one disease to study subjects with a different disease) has yielded interesting observations in other circumstances. Given the well documented association of cannabis use with schizophrenia, we used the summary statistics from a large GWAS study of subjects with cannabis use disorder to study this cohort. To our surprise, PRS based on that GWAS data captured a significant fraction of the variance in risk for Type 2 schizophrenia in the African-Americans in the cohort we are studying (red box in figure 4).

**Figure 4:**
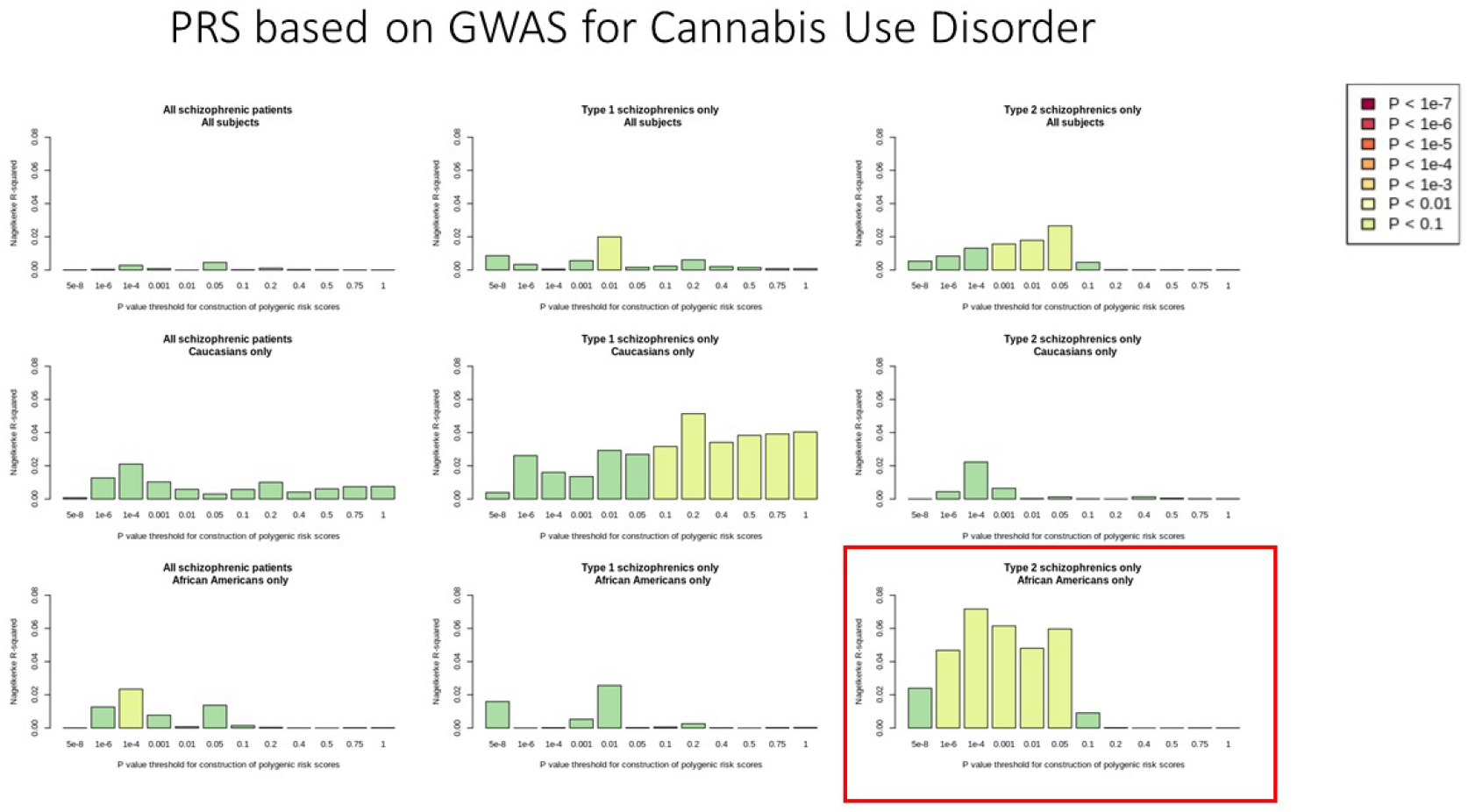
The fraction of variance in risk of schizophrenia captured by PRS for cannabis use disorder (Levey et al. 2023). We do not attempt to explain the complicated pattern revealed by the entire figure. For the purposes of this report, the only significance of this data is that it confirms the biologically reasonable assumption that there are genetic factors which influence the risk of Type 2 schizophrenia in the African-Americans in this cohort in spite of the fact that those genetic factors were not identified in GWAS studies which did not take into account the Type 1 / Type 2 distinction.

We do not attempt to explain the complicated pattern seen in the various panels of figure 4. The relevance of the observation illustrated in figure 4 to this work is ONLY that it confirms that there is indeed a genetic component to the risk of Type 2 schizophrenia in the African-Americans in this cohort in spite of the fact that the risk is not captured in the Psychiatric Genetic Consortium GWAS studies which do not distinguish Type 1 from Type 2 disease.

### A potentially important finding regarding treatment resistant schizophrenia (TRS)

We took advantage of the seminal study by Pardiñas et al. (2022) in which they used a subtractive method and GWAS data from two studies of patients with treatment resistant schizophrenia (TRS) to impute the genetic factors responsible for treatment resistence in schizophrenia. As was made clear in that paper, PRS based on that imputed-GWAS data is not PRS for treatment resistant schizophrenia. It is rather a polygenic score for the genetic risk factors which differentiate TRS from non-TRS.

That study was done using data from exclusively Caucasian cohorts and the completely unexpected result reported here is that PRS for the genetic factors associated with treatment resistance in Caucasian patients accounts for variance in the risk of schizophrenia exclusively in Type 2 patients (red box in figure 5).

**Figure 5:**
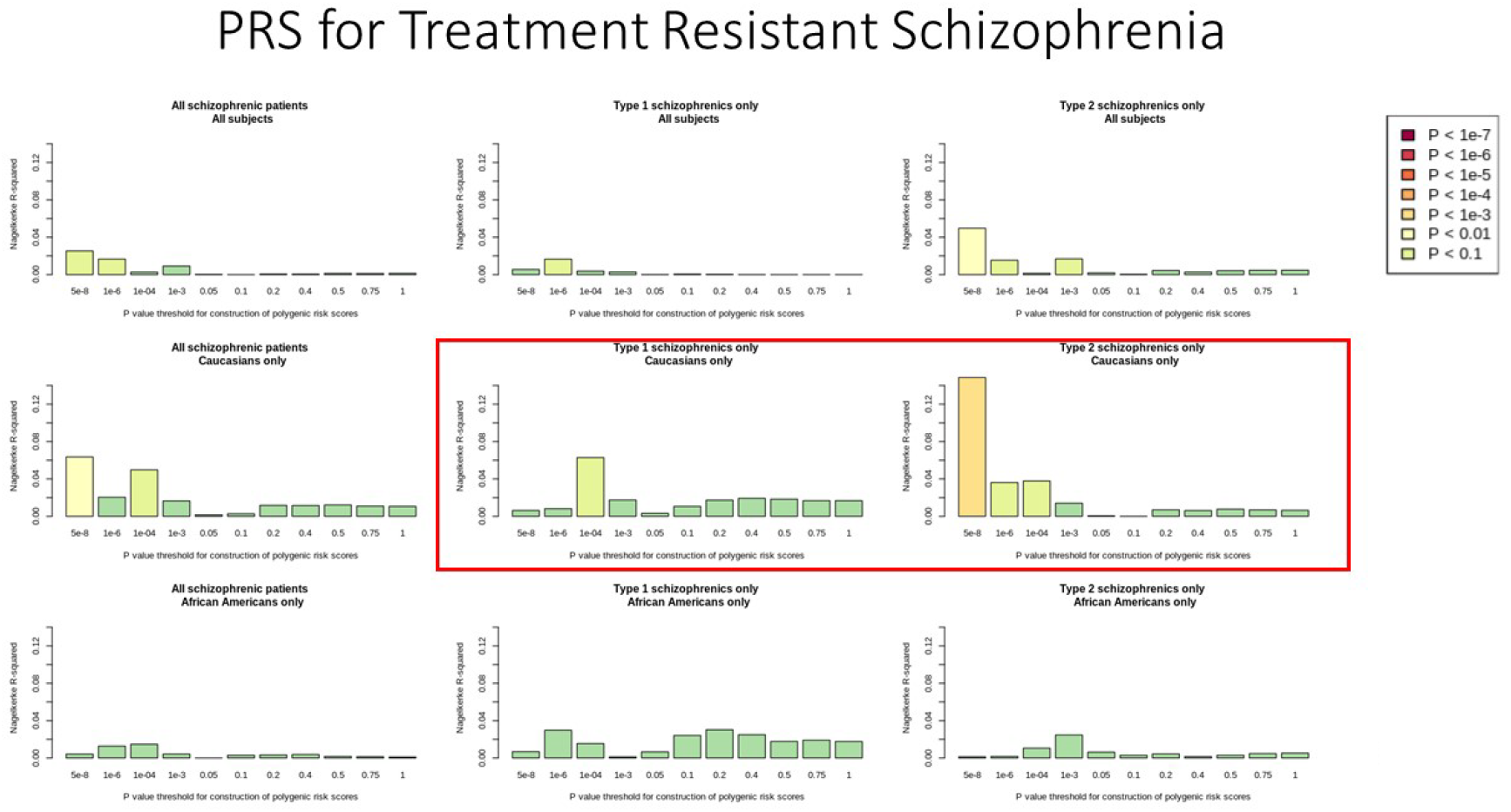
The fraction of variance in risk of schizophrenia captured by PRS for genetic factors responsible for treatment resistance in schizophrenia based on the summary statistics from imputed-GWAS study of Pardiñas et al. (2022)

## Discussion

We have previously described two groups of schizophrenia patients, “Type 1” patients with a relatively normal DLPFC transcriptome and “Type 2” patients with hundreds of differentially expressed genes (Bowen et al. 2019). In this context “relatively normal” means “with very few transcripts whose differential expression survives Bonferroni correction in this cohort”. Using the commonly used FDR criterion there are, of course, many differentially expressed genes in the DLPFC of Type 1 patients and we do not intend to imply that their DLPFC is normal, only that it is very much more normal than that of the Type 2 patients. There are obvious limitations of that study and without some replication of this result based on some completely different molecular study, it is not surprising that Bowen et al. has received very little attention. **We have now shown that Type 1 and Type 2 schizophrenia have dramatically different genetic architectures, a result which very strongly supports the hypothesis that these are two biologically different diseases**.

There are two of important caveats which must be kept in mind when evaluating the results of this study and incorporating them into future scientific studies and clinical trials.

The first caveat is that Type 1 schizophrenic patients do not have a “normal” DLPFC transcriptome. They simple have a transcriptome which is more normal than that of Type 2 patients especially when a Boneferroni correction is used as the criterion for statistical significance. When the much less stringent false discovery rate criterion is applied, both the Type 1 and Type 2 patients have unequivocally abnormal DLPFC transcriptomes.

The second caveat is there is nothing in the results of this study or the previous study (Bowen et al., 2019) which address the question of whether the Type 1 vs Type 2 distinction represents a spectrum or a dichotomy. There are individual patients whose DLPFC transcriptome is more abnormal than that of the typical Type 1 patient, but less abnormal than that of the typical Type 2 patient. And just as the phenotype of dementia may in some patients be due to a combination of comorbid vascular dementia and Alzheimers disease, so there may be patients with schizophrenia due to comorbid Type 1 and Type 2 disease.

An important unexpected finding in this study was the dramatic difference in the genetic architecture of schizophrenia in the Caucasians and African-Americans in this cohort. When interpreting that result it is important to realize that this Medical Examiner derived cohort is almost certainly not representative of the general population. In this particular cohort African-American ancestry may be a surrogate measure of lower socio-economic status and more specifically the African-Americans in the HBCC cohort may have had poorer prenatal obstetric care than the Caucasians in that cohort and/or have been preferentially subject to adverse enviromental influences during early post-natal life.

Althouth there is data in the literature to support those possibilities, this study makes no attempt to evaluate that literature or to test that hypothesis. Never-the-less, both the scientific and the public health implications of this results of the present study are obvious and the observation deserves further study.

Finally, we have made the serndipidous observation that PRS for the genetic factors for the risk of treatment resistance in schizophrenic patients account for variance in the risk of schizophrenia in Type 2, but not Type 1 patients. This does NOT imply that TRS patients are exclusively or even primarily Type 2 patients. TRS is a very complicated phenotype which presumably results from a wide variety of environmental and genetic causes and it would be unrealistic to expect some simple explanation for its underlying biology. More importantly, we have preliminary unpublished data about the demographics of clozapine treatment which suggest that TRS may be equally common in Type 1 and Type 2 patients. This observation does, however, suggest that a study of the genes and genetic polymorphisms driving the PRS effect seen in figure 5 may provide important insights into the biology of both TRS and Type 2 schizophrenia. The implications of this result for efforts to use gene expression data to identify novel pharmacologic therapies for schizophrenia are obvious.

## Data Availability

All data produced in the present study are available upon reasonable request to the authors.

## Footnotes

1 For two excellent tutorials on the use of this software and instructions downloading it see: https://pdhp.isr.umich.edu/wp-content/uploads/2021/03/2021-03-PDHP_Sociogenomics_Part2_Ware.pdf and https://choishingwan.github.io/PRS-Tutorial/prsice/

2 There is a similar problem with the clustering algorithm in Seurat used to cluster single nucleus RNAseq data and identify cell types. It is well recognized that when pressed far enough that algorithm will distinguish subtypes which are not biologically meaningful. To address this problem, Grabski, Street, and Irizarry, 2023 have reported an approach based on the algorithm used here for clustering with single-cell RNA-sequencing data which incorporates significance testing of the clusters.

3 The inverse hyperbolic sine transformation is commonly used by economists (see, for example, Pence 2006) and might have general applications in the biologic sciences.

## Notes

### Competing Interest Statement

The authors have declared no competing interest.

### Funding Statement

This study did not receive any funding.

### Author Declarations

The study used ONLY openly available human data that were are located at the NIMH Data Archive.

### Summary of Updates

PRS are now calculated by the authors using genetic data from NDA rather than being provided by Duncan et al. This allows the calculation of the multiple PRS now being reported rather than only PRS based on the PGC wave 3 core cohort summary data. All of the figures are revised, the Results are revised to include these new PRS, and the Discussion is revised and substantially expanded to include these new results.

